# Family-based genome-wide association study of leprosy in Vietnam

**DOI:** 10.1101/2020.01.30.20018432

**Authors:** Chaima Gzara, Monica Dallmann-Sauer, Marianna Orlova, Nguyen Van Thuc, Vu Hong Thai, Vinicius M. Fava, Marie-Thérèse Bihoreau, Anne Boland, Laurent Abel, Alexandre Alcaïs, Erwin Schurr, Aurélie Cobat

## Abstract

Leprosy is a chronic infectious disease of the skin and peripheral nerves with a strong genetic predisposition. Recent genome-wide approaches have identified numerous common variants associated with leprosy, almost all in the Chinese population. We conducted the first family-based genome-wide association study of leprosy in 622 affected offspring from Vietnam, followed by replication in an independent sample of 1189 leprosy cases and 671 controls of the same ethnic origin. The most significant results were observed within the HLA region, in which six SNPs displayed genome-wide significant associations, all of which were replicated in the independent case/control sample. We investigated the signal in the HLA region in more detail, by conducting a multivariate analysis on the case/control sample of 319 GWAS-suggestive HLA hits for which evidence for replication was obtained. We identified three independently associated SNPs, two located in the HLA class I region (rs1265048: OR=0.69 [0.58-0.80], combined *p*-value = 5.53×10^−11^; and rs114598080: OR=1.47 [1.46-1.48], combined *p*-value = 8.77×10^−13^), and one located in the HLA class II region (rs3187964 (OR=1.67 [1.55-1.80], combined *p*-value = 8.35×10^−16^). We also validated two previously identified risk factors for leprosy: the missense variant rs3764147 in the *LACC1* gene (OR=1.52 [1.41-1.63], combined *p*-value = 5.06×10^−14^), and the intergenic variant rs6871626 located close to the *IL12B* gene (OR=0.73 [0.61-0.84], combined *p*-value = 6.44×10^−8^). These results shed new light on the genetic control of leprosy, by dissecting the influence of HLA SNPs, and validating the independent role of two additional variants in a large Vietnamese sample.

## Introduction

Leprosy is a chronic infectious disease caused by *Mycobacterium leprae*. It primarily affects the skin and peripheral nerves, and can cause an irreversible impairment of nerve function, often leading to severe disabilities and social stigma. Despite a decrease in its prevalence over the last two decades, leprosy remains a major public health problem in regions of endemic countries, with over 200,000 new cases detected in 2018 (https://www.who.int/gho/neglected_diseases/leprosy/en/). However, this number of cases is probably a severe underestimate of the true incidence (1). The clinical disease develops in a minority of exposed individuals, manifesting as a spectrum of disease symptoms reflecting the interactions between the host immune response and the bacterium (2). Tuberculoid and lepromatous leprosy are at opposite ends of the clinical spectrum and are associated with a relatively stable host immune status while borderline categories of the disease are associated with an unstable immune response to the bacilli.

Only a subset of individuals develops clinical leprosy after sustained exposure to *M. leprae*. Due to the extreme reductive evolution of the *M. leprae* genome (3), it is highly unlikely that differences in susceptibility or clinical manifestations are governed by the *M. leprae* strain or by intrastrain variation (4). From the early observations of familial clusters of leprosy cases to recent whole-exome sequencing studies identifying genetic variants associated with leprosy, there is strong evidence to suggest that the development of this disease is under tight human genetic control (2, 5). Genetic studies, including positional cloning analyses (6-8), genome-wide association studies (9-15), and, more recently, whole-exome sequencing (16) have identified several susceptibility loci for leprosy (reviewed in (17) and (5)), and have demonstrated the involvement of both innate and adaptive immune responses in this disease (18). Almost all the susceptibility loci identified in genome-wide studies to date were found in the Chinese population. The association with leprosy has been replicated in the Vietnamese population for some of these loci (19, 20), but others were found to be associated with type-1 reactions (T1Rs), a pathological inflammatory response afflicting a subgroup of leprosy patients and resulting in peripheral nerve damage (21-23), rather than with leprosy itself. Here, we performed a two-stage family-based genome-wide association study on leprosy *per se*, i.e. leprosy independent of its clinical subtype, in a large collection of Vietnamese families including 622 affected offspring, followed by a replication study in an independent sample of 1189 leprosy cases and 671 controls.

## Materials and Methods

### Ethics statement

The study was conducted in accordance with the Declaration of Helsinki. Written informed consent was obtained from all adult subjects participating in the study. All minors agreed to take part, and a parent or guardian provided informed consent on their behalf. The study was approved by the Research Ethics Board at the Research Institute at McGill University Health Centre in Montreal (REC98-041) and the regulatory authorities and ethics committees of Ho Chi Minh City in Vietnam (So3813/UB-VX and 4933/UBND-VX).

### Samples and study design

Over twenty-five years (1990-2015), we have, in close collaboration with the Dermatology and Venereal Diseases Hospital of Ho-Chi-Minh City (Vietnam), recruited a large sample of nuclear families with at least one child diagnosed with leprosy and including either both parents or unaffected sibling(s) if one of the parents was unavailable. Leprosy was diagnosed by at least two independent and experienced physicians and was classified into MB or PB subtype according to the operational WHO-96 definition based solely on the number of lesions (2). For replication purposes, we collected an independent case/control sample of Vietnamese origin. The same criteria were used for leprosy diagnosis as in the family-based discovery study. The controls had no personal or family (among first-, second-or third-degree relatives) history of leprosy or tuberculosis.

### GWAS genotyping and imputation

The genotypes of all children and parents (or siblings) were determined with the Illumina Human 660w Quad v1 bead chip containing 592,633 single-nucleotide polymorphisms (SNPs) during the discovery phase. Genotypes were called with the Illuminus algorithm (24). Quality control (QC) was performed on genotype data with PLINK software (25) and the “GASTON” package in R (https://CRAN.R-project.org/package=gaston). Only autosomal SNPs were considered. SNPs with a call-rate < 0.95, more than 10 Mendelian errors, a minor allele frequency (MAF) < 0.05 or displaying significant departure from Hardy-Weinberg equilibrium (*p* < 10^−5^) were removed from the analyses, resulting in a final set of 412,639 high-quality autosomal SNPs. Individuals with a call-rate < 0.95, more than 10 Mendelian errors and a heterozygosity rate more than three standard deviations on either side of the mean were excluded. In total, 1850 individuals were genotyped and 1749 fulfilled the quality control criteria, including 622 leprosy-affected offspring, from 481 informative families (Table S1).

Before imputation in the discovery sample, A/T or G/C SNPs were removed to prevent strand mismatches between the study sample and the reference sample used for imputation. The remaining high-quality SNPs were pre-phased with SHAPEIT (26) v2 software using to the duoHMM method, to incorporate the known pedigree information and improve phasing (27). Genotypes from the 1000 Genomes Project phase 3 haplotype reference panel released in October 2014 were further imputed with IMPUTE2 software (28). An information imputation threshold (Info) > 0.9 was applied to capture most of the common variants (MAF > 5%) with reasonable confidence, leading to the retention of ∼5 million additional variants (SNPs and INDELs).

### Replication strategy

We used three different strategies to select SNPs for genotyping in the replication sample, according to the type of variant. First, for genome-wide suggestive (*p*-value < 10^−5^) hits located outside the HLA region (non-HLA SNPs), we used a linkage disequilibrium (LD)-based clumping procedure, implemented in PLINK 1.9 (25, 29), to identify independent loci. This procedure considers all SNPs with *p*-values for association below 10^−5^, and forms clumps of these ‘index’ SNPs, together with all other SNPs in LD with them at an r^2^ threshold of 0.5 and in close physical proximity (less than 5 Mb away from the index SNPs). Only SNPs with a *p*-value below a secondary significance threshold of 10^−4^ are clumped. As input for the clumping procedure, we used association *p*-values from the discovery GWAS and LD patterns estimated from the Kinh in Ho Chi Minh City, Vietnam (KHV) population of the 1000 Genomes phase 3 reference set. We selected one SNP from each clump for genotyping in the replication sample, based on technical considerations. We applied a second strategy specifically to the non-HLA SNPs previously associated with leprosy at a genome-wide level of significance (Table S2). For these SNPs, we used a 10^−2^ *p*-value threshold to select the SNPs for the replication study. Finally, for SNPs located within the HLA region, we used a third strategy, because there were large numbers of SNPs displaying suggestive evidence of association, including genome-wide significant (*p*-value < 5×10^−8^) results, and complex LD patterns. We selected a set of 88 SNPs located at positions from 30 Mb to 33 Mb on chromosome 6, on the basis of preliminary association results, LD structure and technical considerations for genotyping, and we genotyped these SNPs in the replication sample. We then used these SNPs to impute, in the replication sample, all the variants for which there was genome-wide suggestive evidence of association.

For the replication sample, genotypes were determined by FLUIDIGM targeted sequencing or with SEQUENOM technology. For FLUIDIGM data, we used the Genome Analysis Toolkit (30) (GATK v3.3) to process the bam files. Individual genomic variant call files (gVCF) were generated with GATK HaplotypeCaller, and joint genotyping was performed with GATK GenotypeGVCFs. SNPs with a call-rate < 0.95 or significant departure from Hardy-Weinberg equilibrium (*p* < 0.001) were removed from the analyses. Individuals with a call-rate < 0.95 were excluded. Imputation in the MHC region was performed with SHAPEIT v2 (26) and IMPUTE2 (28) software and the 1000 Genomes Project phase 3 haplotype reference panel.

### Statistical methods

In the discovery phase, a family-based association test was used to estimate the non-random transmission of alleles from heterozygous parents to leprosy-affected offspring. The analysis was carried out under the additive model, with FBATdosage v2.6, for genotyped and imputed variants (19). Alleles for which some evidence of association was obtained were also analyzed by conditional logistic regression, as previously described (31-33). Briefly, for each affected child, up to three matched unaffected pseudosibs were formed by all other possible combinations of parental alleles. The real affected offspring were compared with the pseudosibs in a matched case-control design. This approach generated odds ratio estimates and made it possible to perform multivariate stepwise regression. Conditional logistic regression was also used for a combined analysis of the discovery family and replication case-control samples. In this combined analysis, all cases and controls from the case-control study were grouped together in a stratum for analysis, in combination with the strata consisting of the affected offspring and their pseudosibs, as previously described (32, 33). Conditional logistic analysis was performed with the LOGISTIC procedure implemented in SAS software v. 9.3 (SAS Institute, Cary, North Carolina, USA).

Case/control association analyses of genotyped and imputed SNPs in the replication sample were performed by logistic regression analysis, under the additive model, with SNPTEST software (28, 34). A one-tailed test was performed with the alternative hypothesis that the leprosy susceptibility alleles of the discovery sample were also susceptibility factors in the replication sample, and a *p*-value threshold of 0.01 was used in this one-tailed test for the detection of significant replication. We also performed a multivariable analysis of the HLA SNPs replicated in the case/control replication sample, using the best subset selection method of the SAS 9.3 logistic regression procedure.

## RESULTS

### Genome-Wide Analyses in the Primary Cohort

We carried out a family-based GWAS on leprosy *per se* in 481 informative families consisting of 1749 individuals, including 622 offspring with leprosy (Table S1). For the discovery phase, 5,533,693 high-quality genotyped and imputed autosomal variants were analyzed. Principal component analysis of our samples with data for individuals from the 1000 Genomes database revealed clustering with the 1000 Genomes KHV population (Figure S1). A Manhattan plot of the association with leprosy *per se* is shown in Figure 1. The most significant results were obtained for the HLA region, in which genome-wide significant association with leprosy was detected for six SNPs (*p*-value < 5×10^−8^) (Table 1), and genome-wide suggestive association with leprosy was detected for 358 SNPs (*p*-value < 10^−5^). Outside the HLA region, there was no significant deviation from expectations on the quantile-quantile plot of the GWAS results (Figure S2). In total, 21 SNPs from eight independent clusters (Table S2) on chromosomes 4, 5, 9, 10, 11, 14 and 21 (2 clusters) displayed genome-wide suggestive association with leprosy (*p*-value < 10^−5^). Finally, evidence of association was found in the Vietnamese discovery sample, for five of the 33 genome-wide significant previously published non-HLA hits, with a *p*-value threshold of 10^−2^ and the same risk allele as in the original study (Table S3).

**Table 1.** Association statistics in the discovery and replication samples for the six genome-wide significant HLA SNPs and the five eviously published non-HLA SNPs replicated in the discovery GWAS

| SNP | Chr | Position<sup>\$</sup> | m/M | Discovery | | | | Replication | | |
| --- | --- | --- | --- | --- | --- | --- | --- | --- | --- | --- |
|  |  |  |  | Type | MAF | OR* [95% CI] | P | MAF | P<sup>#</sup> | OR*<sup>#</sup> [95% CI] |
| Significant HLA hits |  |  |  |  |  |  |  |  |  |  |
| rs3095309 | 6 | 31091475 | T/C | imputed | 0.476 | 1.6 [1.43-1.77] | 8.2x10<sup>-09</sup> | 0.44 | 9.46x10<sup>-05</sup> | 1.30 [1.16 - Inf[ |
| rs3094194 | 6 | 31136240 | G/A | imputed | 0.46 | 1.71 [1.53-1.88] | 1.4x10<sup>-08</sup> | 0.44 | 5.03x10<sup>-06</sup> | 1.36 [1.21 - Inf[ |
| rs2844633 | 6 | 31096189 | T/C | imputed | 0.269 | 0.56 [0.38-0.75] | 1.7x10<sup>-08</sup> | 0.30 | 4.20x10<sup>-04</sup> | 0.79 ]0 - 0.89] |
| rs2844634 | 6 | 31096184 | C/G | imputed | 0.269 | 0.56 [0.38-0.75] | 1.7x10<sup>-08</sup> | 0.30 | 4.65x10<sup>-04</sup> | 0.79 ]0 - 0.89] |
| rs2394945 | 6 | 31220971 | G/C | imputed | 0.353 | 1.73 [1.54-1.93] | 4.2x10<sup>-08</sup> | 0.35 | 9.20x10<sup>-07</sup> | 1.42 [1.25 - Inf[ |
| rs3094196 | 6 | 31093947 | G/A | imputed | 0.269 | 0.57 [0.38-0.75] | 4.9x10<sup>-08</sup> | 0.30 | 4.70x10<sup>-04</sup> | 0.79 ]0 - 0.89] |
| Previously published non-HLA hits replicated in the discovery GWAS |  |  |  |  |  |  |  |  |  |  |
| rs3762318 | 1 | 67597119 | G/A | genotyped | 0.05 | 0.52 [0.12-0.93] | 4.41x10<sup>-03</sup> | 0.06 | 1.57x10<sup>-01</sup> | 0.86 ]0 - 1.10] |
| rs2221593 | 1 | 212873431 | T/C | genotyped | 0.229 | 1.33 [1.13-1.53] | 1.94x10<sup>-04</sup> | Failed call rate < 95% |  |  |
| rs2058660 | 2 | 103054449 | G/A | imputed | 0.414 | 0.75 [0.57-0.92] | 5.66x10<sup>-03</sup> | 0.40 | 6.46x10<sup>-02</sup> | 0.90 ]0 - 1.01] |
| rs6871626 | 5 | 158826792 | A/C | imputed<sup>£</sup> | 0.324 | 0.71 [0.58 - 0.86] | 5.12x10<sup>-04</sup> | 0.32 | 1.37x10<sup>-05</sup> | 0.74 ]0 - 0.83] |
| rs3764147 | 13 | 44457925 | G/A | genotyped | 0.396 | 1.41 [1.18 - 1.67] | 6.56x10<sup>-05</sup> | 0.39 | 2.36x10<sup>-11</sup> | 1.61 [1.43 - Inf[ |

m/M = minor/Major alleles; MAF = minor allele frequency; OR = odds ratio; CI = confidence interval
<sup>\$</sup> positions are given according to the hg19 genome build
\* with respect to the minor allele
<sup>#</sup> one-tailed test
<sup>£</sup> imputed using the 1000 Genomes phase 1 reference panel
Bold typeface indicates SNPs significant in the replication sample

**Figure 1.**
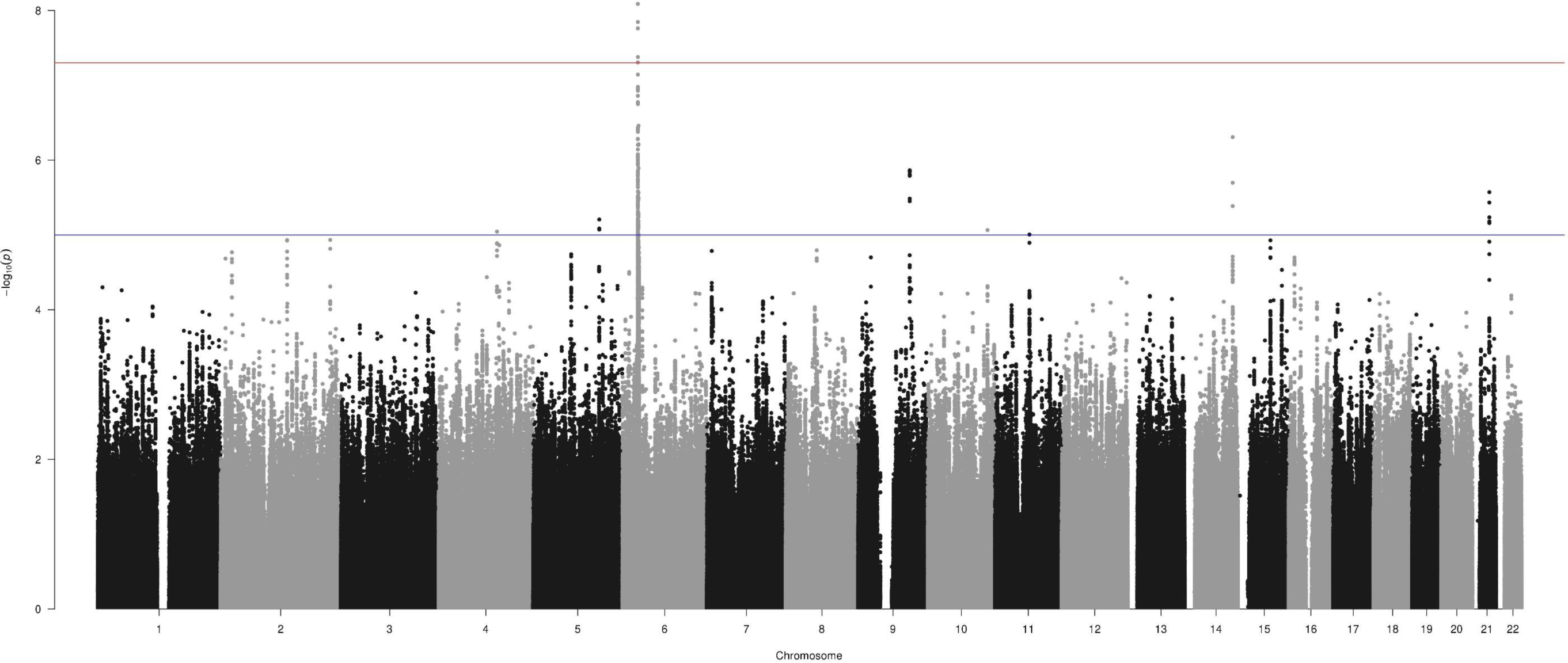
Manhattan plot of the results for association with leprosy *per se* in the family-based GWAS. Manhattan plot showing results of the family-based genome-wide association study of leprosy *per se* in 622 affected offspring, for 5,533,693 variants (minor allele frequency > 5% and info > 0.8). The –log_10_(*p-*value) for each variant (*y-*axis) is plotted according to its chromosomal position (*x-*axis, build hg19). The red and blue lines indicate the genome-wide significance (5.0×10^−8^) and suggestive (10^−5^) thresholds, respectively.

### Replication Study

#### Outside the MHC region

Within each of the 8 loci for which suggestive association with leprosy was detected, we genotyped one SNP in an independent replication cohort of 1189 leprosy cases and 674 controls. We also genotyped the 5 previously published non-HLA hits replicated in the discovery cohort with a *p*-value threshold of 10^−2^. In total, 10 of the 13 SNPs satisfying the quality control filters in the replication cohort were tested for association in the 1181 cases and 668 controls. No evidence of replication was obtained for any of the suggestive GWAS hits that could be tested in the replication sample (Table S2). For two suggestive GWAS hits, on chromosomal region 4q26 and 9q31.1, the genotyping failed in the replication sample. Replication was observed for two previously published hits fulfilling the genotyping quality criteria (Table 1 and Table S3). The first, rs3764147, located on chromosome 13, is a missense variant (p.Ile254Val) of the *LACC1* gene with a highly significant genome-wide *p*-value in the combined analysis of the discovery family and the replication case/control samples (*p* = 5.1×10^−14^). The odds ratio (OR) for developing leprosy for AG subjects vs. GG subjects (or AA vs. AG) was 1.52 [95% confidence interval: 1.41-1.63] (Table 2). This SNP belongs to a bin of 10 SNPs in strong LD (r^2^ > 0.8) in the 1000 Genomes KHV population that are located either within or just downstream of the *LACC1* gene (Table S4). The second replicated hit, rs6871626, is located on chromosome 5, 69 kb 5’ to *IL12B*. A borderline non-significant genome-wide *p*-value of 6.4×10^−8^ was obtained for this SNP, with an OR of developing leprosy for AC vs. AA (or CC vs. AC) of 0.73 [0.61-0.84]. This SNP is not part of the 1000 Genomes phase 3 reference panel and no LD information was available for the KHV population, but it was successfully sequenced in the genome part of The Genome Aggregation Database (gnomAD) (35).

**Table 2.** Association statistics in the discovery GWAS, the case/control replication and the combined GWAS and replication samples for five independent SNPs identified by multivariate analysis

| SNP<br>(chr:position<sup>\$</sup>) | m/M | Gene | MAF<sup>@</sup> | Discovery GWAS | | Replication | | Combined GWAS and<br>replication sample | |
| --- | --- | --- | --- | --- | --- | --- | --- | --- | --- |
|  |  |  |  | *P* value | OR* [95% CI] | *P* value<sup>#</sup> | OR** [95% CI] | *P* value | OR* [95% CI] |
| rs1265048<br>(6:31081409) | C/T | *C6orf15* | 0.49 | 8.72x10<sup>-06</sup> | 0.63 [0.52 -0.63] | 1.58x10<sup>-06</sup> | 0.72 ]0 - 0.81] | 5.53x10<sup>-11</sup> | 0.69 [0.58-0.80] |
| rs114598080<br>(6:31255442) | A/G | *HLA-C* | 0.10 | 3.90x10<sup>-06</sup> | 1.85 [1.39 -2.44] | 4.39x10<sup>-11</sup> | 2.06 [1.7 - Inf[ | 8.77x10<sup>-13</sup> | 1.47 [1.46-1.48] |
| rs3187964<br>(6:32605207) | C/G | *HLA-DQA1* | 0.41 | 1.28x10<sup>-06</sup> | 1.52 [1.25 - 1.85] | 5.28x10<sup>-13</sup> | 1.72 [1.51 - Inf[ | 8.35x10<sup>-16</sup> | 1.67 [1.55-1.80] |
| rs3764147<br>(13:44457925) | G/A | *LACC1* | 0.35 | 6.56x10<sup>-05</sup> | 1.41 [1.18 - 1.67] | 2.36x10<sup>-11</sup> | 1.61 [1.49-Inf[ | 5.06x10<sup>-14</sup> | 1.52 [1.41-1.63] |
| rs6871626<sup>£</sup><br>(5:158826792) | A/C | *IL12B* | 0.35 | 5.12x10<sup>-04</sup> | 0.71 [0.58 - 0.86] | 1.37x10<sup>-05</sup> | 0.74 ]0-0.86] | 6.44x10<sup>-08</sup> | 0.73 [0.61-0.84] |

m/M = minor/Major alleles; MAF = minor allele frequency; OR = odds ratio; CI = confidence interval
<sup>\$</sup> positions are given according to the hg19 genome build
<sup>@</sup> MAF was estimated from the 939 leprosy unaffected parents of the discovery GWAS and 668 controls of the replication sample
\* with respect to the minor allele
<sup>#</sup> one-tailed test
<sup>£</sup> imputed using the 1000 Genomes phase 1 reference panel

#### Within the MHC region

We genotyped 88 SNPs within the MHC region, and used them to impute the 364 SNPs with a discovery *p*-value < 10^−5^. More than 98% of these SNPs (358/364) were imputed with an information score > 0.8. The six HLA SNPs with genome-wide significance were replicated, all with a one-tailed *p*-value < 5×10^−4^ (Table 1). In addition, 319 of the 358 genome-wide suggestive HLA SNPs were replicated, with a one-tailed *p*-value threshold of 0.01. They clustered in two intervals, from 31 Mb to 31.5 Mb and from 32.5 Mb to 32.7 Mb (Figure 2, Table S5). We further investigated the number of SNPs in the HLA region required to capture the full association signal, by performing a multivariable analysis. This analysis was conducted in the replication sample, which provided the most reliable set of SNPs and facilitated the analysis because of its case/control nature. Before performing this multivariable analysis, we clumped the SNPs, as described in the methods section, on the basis of their genotyping status in the replication sample (genotyped SNPs were favored over imputed ones), their one-tailed replication *p*-value and a LD r^2^ threshold of 0.5.

**Figure 2.**
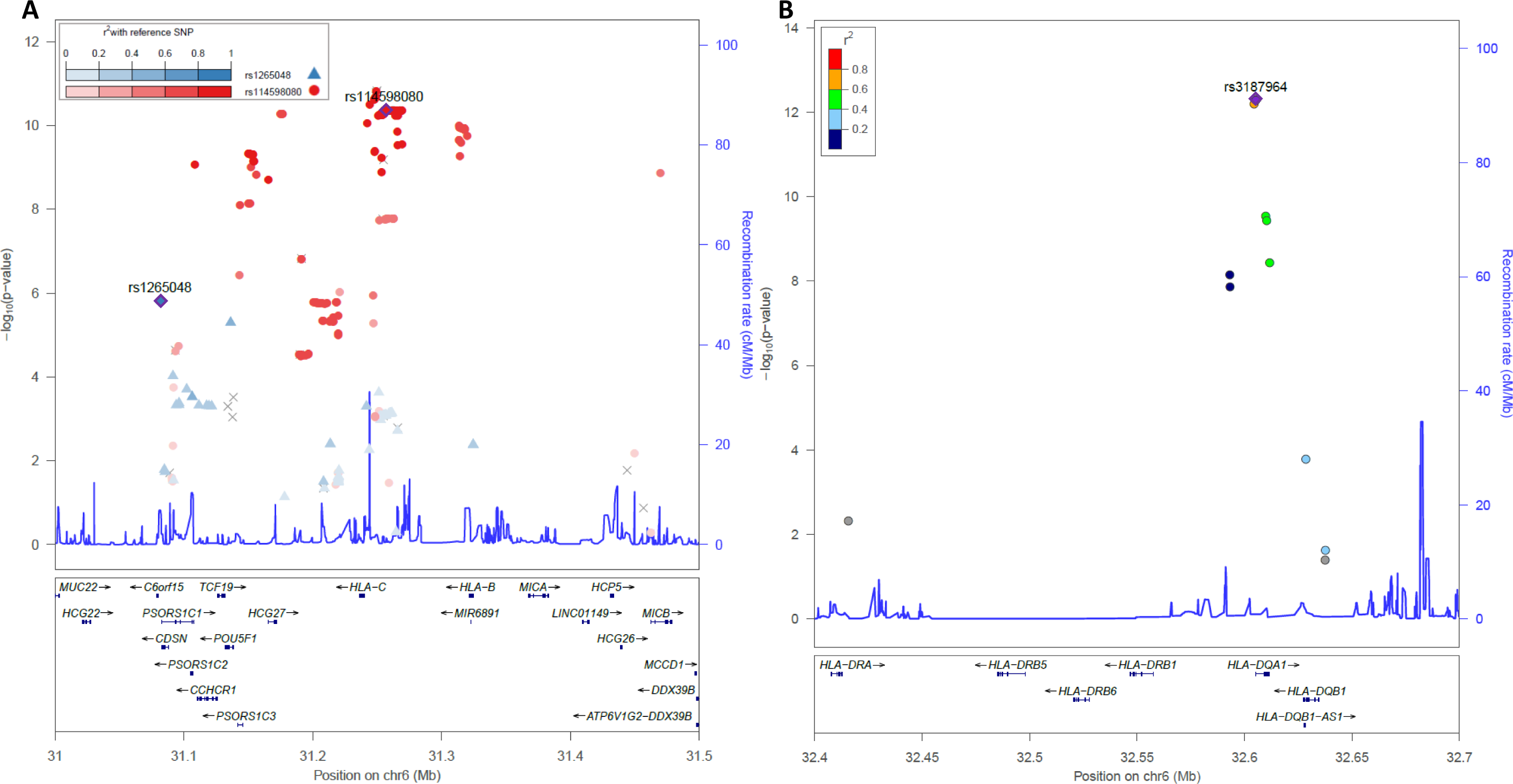
Regional association plot for HLA class I (A) and class II (B) SNPs in the case/control replication sample. Locus zoom plot showing one-tailed *p*-values for association within the HLA region. The three independent SNPs identified by multivariate analysis, rs1265048 (A), rs114598080 (A) and rs3187964 (B), are represented as blue, red and purple diamonds, respectively. The colors indicate the pairwise linkage disequilibrium (*r*^2^) with the three independent SNPs, as calculated in the 1000 Genomes KHV population. SNPs not found in the reference populations are shown in gray.

The clumping procedure identified 18 index SNPs with one-tailed *p*-value < 0.01, which were then tested in the multivariable model with the best subset method (SCORE) implemented in the logistic regression procedure of the SAS software (Table S6). We found that three SNPs (rs3187964, rs114598080 and rs1265048) were required to capture the full association signal in the HLA region (Table S7). The addition of a fourth SNP did not significantly improve the multivariable model (*p*-value = 0.23). The most significant SNP in the multivariable model was rs3187964 (multivariable *p*-value = 7.8×10^−8^). The strength of association was much lower for rs114598080 (multivariable *p*-value = 3.75×10^−5^), and even lower for rs1265048, (multivariable *p*-value = 0.011). These three SNPs were sufficient to account for the HLA association signal found in our GWAS.

We then investigated in more detail the location, LD in the 1000 Genome KHV population and univariate results in the combined sample of these three HLA SNPs (Table 2). The rs3187964 SNP belongs to the HLA interval corresponding to the HLA class II region (Figure 2B). In the combined analysis of the discovery and replication samples, it gave a univariate *p*-value of 8.3×10^−16^ with an OR for developing leprosy for CG vs. GG (or CC vs. CG) of 1.67 [1.55-1.80]. This SNP is located in the 5’ untranslated region of the *HLA-DQA1* gene and was not in strong LD (r^2^ > 0.8) with other SNPs in the 1000 Genomes KHV population in an LDproxy analysis (36). The other two SNPs are in the first HLA interval, which includes HLA class I genes, together with many other genes (Figure 2A). The rs114598080 SNP, located in a CTCF binding site (ENSR00000262841) 15.6 kb upstream of the *HLA-C* gene, gave a univariate *p*-value of 8.8×10^−13^ in the combined analysis, with an OR for developing leprosy for AG vs. GG (or AA vs. AG) of 1.47 [1.46-1.48]. This SNP belongs to a bin of 245 SNPs in strong LD that span almost 200 kb (Table S8). The third SNP, rs1265048, is located near the *C6orf15* (1.07 kb), *PSORS1C1* (1.12 kb), and *CDSN* (1.2 kb) genes and it gave a univariate *p*-value of 5.5×10^−11^ in the combined analysis, with an OR for developing leprosy for CT vs. TT (or CC vs. CT) of 0.69 [0.58-0.80]. It was not in strong LD with other SNPs in the 1000 Genomes KHV population in LDproxy analysis.

### Discussion

We performed the first family-based GWAS for leprosy in a Vietnamese population. The main susceptibility loci identified were in the HLA region. The most significant SNP, rs3187964, is located in the MHC class II region, in the 5’ untranslated region of *HLA-DQA1* and did not display strong LD (r^2^ > 0.8) with other SNPs in the region in the 1000 Genomes KHV population. Several studies have consistently reported the involvement of HLA class II alleles, mostly *HLA-DRB1* alleles, as key genetic factors controlling susceptibility to various forms of leprosy (11, 18, 37-39). A recent HLA imputation-based meta-analysis in the Chinese population identified *HLA-DRB1* and *HLA-DQA1* alleles and specific amino acids of HLA-DRβ1 as independent protective factors for leprosy (37). Interestingly, in the GTex project V7 (https://gtexportal.org/home/), rs3187964 was identified as a strong eQTL for several HLA class II genes with *p*-values below 10^−50^ for *HLA-DQB1, HLA-DQB2, HLA-DQA1* and *HLA-DQA2* in multiple tissues, including skin and nerves (Table S9). It was also found to be a splice QTL for *HLA-DQA1* (nominal *p*-value = 3.3×10^−7^, FDR = 0.000155). HLA-DQ genes have been implicated in several autoimmune and inflammatory diseases (40) and, recently, in susceptibility to streptococcal disease (41).

The other two independent SNPs associated with leprosy in the HLA region are located in the class I region. The most significant, rs114598080, belongs to a large bin of SNPs in strong LD spanning the HLA class I genes *HLA-C* and *HLA-B* and the non-HLA genes *CCHCR1, TCF19, POU5F1, PSORS1C3*, and *HCG27*. The complex LD structure makes it difficult to identify the gene related to this association hit. Nevertheless, one SNP of the bin, rs2394885, was shown to tag the *HLA-C*15:05* allele in the Vietnamese population (42). This SNP was also validated as a leprosy susceptibility locus in an Indian population (42). In the Chinese HLA imputation-based meta-analysis, *HLA-C*08:01* and specific *HLA-C* amino acids were found to be independent protective and risk factors for leprosy (37). The rs114598080 SNP is also in strong LD with a possibly damaging missense variant of the *CCHCR1* gene. CCHCR1 is a protein of unknown function that was recently found to have properties typical of P body-resident proteins (43). In addition, rs114598080 is a strong eQTL in sun-exposed skin and tibial artery for *POU5F1* (p < 10^−10^), also known as *OCT4*, which encodes a master transcription factor for pluripotent cell self-renewal (44) (Table S10). The second SNP of the HLA class I region, rs1265048, is located 1.1 kb upstream of *C6orf15*, 1.1 kb upstream of *PSORS1C1* and 1.2 kb upstream of *CDSN*. Interestingly, the SNP correlates with the expression of several genes, including *HLA-C* (minimum *p*-value = 1.3×10^−5^), and *POU5F1* (minimum *p*-value = 2.8×10^−5^) in particular, sporadically in some tissues (Table S11).

In addition to the HLA locus, we were able to consistently replicate the *LACC1* and *IL12B* loci previously associated with leprosy in the Chinese population both in the discovery GWAS and the replication sample. The *LACC1* leprosy susceptibility SNP rs3764147 is a missense variant (p.Ile254Val) first identified by a leprosy GWAS in the Han Chinese population (9) and then replicated in Indian, African (45), Vietnamese (20), Yi (46), Wenshan and Yuxi (16) Chinese populations. The leprosy susceptibility allele was also shown to be associated with an increased susceptibility to Crohn’s (47, 48) and inflammatory bowel diseases (49, 50). SNP rs3764147 has a PHRED-scaled CADD score (51, 52) of 16, placing it in the top 2.5% of deleterious variants relative to all possible reference genome single-nucleotide variants. Interestingly, recent functional studies have shown that *LACC1* encodes a central regulator of the metabolic function and bioenergetic state of macrophages (53). Furthermore, the rs3764147 G risk allele is associated with lower levels of innate receptor-induced activity in primary human monocyte-derived macrophages (54), potentially accounting for the higher susceptibility to intestinal inflammatory diseases and leprosy.

SNP rs6871626 is located in an intergenic region on chromosome 5. In a previous genome-wide meta-analysis study of ulcerative colitis, the most likely candidate gene assigned to this SNP by Gene Relationship Across Implicated Loci (GRAIL) pathway analysis was *IL12B* (55). GRAIL is a statistical tool that assesses the degree of relatedness of a list of disease regions by text mining PubMed abstracts, for the annotation of candidate genes for disease risk (56). *IL12B* encodes IL-12p40, which is common to two interleukins, IL-12 and IL-23. Both IL-12- and IL-23-dependent signaling pathways play critical roles in human antimycobacterial immunity, through the induction of IFN-γ (57, 58). Autosomal recessive complete *IL12B* deficiency is a genetic etiology of Mendelian susceptibility to mycobacterial diseases (MSMD), a rare condition characterized by predisposition to clinical disease caused by weakly virulent mycobacteria, such as BCG vaccines and environmental mycobacteria, in otherwise healthy individuals (59). The rs6871626 C allele is associated with a risk of leprosy, but has been shown to be protective against ulcerative colitis (55), ankylosing spondylitis and Crohn’s disease (60), inflammatory bowel disease (49) and Takayasu arteritis (61). Interestingly, in patients with Takayasu arteritis, the rs6871626 C allele, which confers susceptibility to leprosy, was associated with abnormally low levels of IL-12 (composed of IL-12p40 and IL-12p35) in plasma and low levels of both IL-12p40 and IL12 in the culture supernatants of patient monocytes/macrophages stimulated with LPS (62).

In conclusion, we performed the first family-based GWAS of leprosy *per se* in the Vietnamese population. The strongest association signals were observed for SNPs within the HLA region. We also validated the independent impact of two additional variants, one in *LACC1* and the second close to *IL12B*, recently reported to play functional roles. We further broke down the association signal within the HLA region into three independent signals, two mapping to the HLA class I region, and one, the leading signal, located in the class II region close to *HLA-DQA1*. More refined analyses based on direct HLA typing, which is lacking so far, are required to characterize further the precise contribution of HLA class I and class II alleles to leprosy susceptibility.

## Data Availability

Data will be available upon request once the manuscript is published in a peer-reviewed journal

## Acknowledgments

We thank all leprosy patients and their family who participated in this study. We thank the staff of leprosy control program in Southern Viêt Nam and the members of leprosy control program at Hospital of D-V of Ho Chi Minh City, Viêt Nam. We thank members of the laboratory of human genetics of infectious diseases and the Erwin Schurr laboratory for helpful discussions. We thank the French national genotyping center (CNG) and Genome Quebec for performing genotyping. This work was supported by grants from the Canadian Institutes of Health Research (CIHR; FDN-143332) to ES, MALTALEP from l’Ordre de Malte to AA and ES, the Agence Nationale de la Recherche (ANR) to AA.

